# Deep Learning and Machine Learning for Early Detection of Alzheimer’s Disease: A Systematic Review and Meta-Analysis

**DOI:** 10.64898/2026.05.21.26353815

**Authors:** Saketh Machiraju

**Affiliations:** Univeristy of California - Santa Cruz, Santa Cruz, CA

**Keywords:** Alzheimer’s disease, machine learning, deep learning, neuroimaging, meta-analysis, diagnostic accuracy, artificial intelligence, mild cognitive impairment, AUC-ROC, multimodal diagnosis

## Abstract

Alzheimer’s disease is a progressive neurodegenerative disorder that poses a growing global public health challenge. Early and accurate diagnosis is critical for effective treatment, clinical trial participation, and disease management. This systematic review and meta-analysis evaluates the diagnostic performance of machine learning (ML) and deep learning (DL) algorithms for detecting Alzheimer’s disease (AD) and mild cognitive impairment (MCI) using neuroimaging and clinical data. Relevant studies were identified from PubMed, IEEE Xplore, and arXiv (2015–2025). Random-effects models were applied to estimate pooled performance metrics (AUC, sensitivity, specificity, and F1-score), and subgroup analyses compared results by model type, imaging modality, and validation strategy. Thirty studies met inclusion criteria, including different diagnosis methods, datasets, and model architectures. The pooled area under the receiver operating characteristic curve (AUC) was 0.962, indicating high overall discriminative accuracy. However, studies relying solely on internal validation or with smaller datasets using pre-processing techniques often reported inflated metrics, suggesting potential overfitting and optimism bias. In summary, ML and DL methods demonstrate strong potential for early AD detection, but standardized evaluation protocols and thorough external validation testing are necessary for real-world clinical translation and adoption.

## 1 Introduction

Recent machine learning (ML) and deep learning (DL) methods can learn subtle disease patterns from structural MRI, PET, and CSF, with growing work in EEG, blood biomarkers, and retinal imaging [1, 2, 3, 4, 5, 6, 7, 8]. In controlled settings, these models sometimes match or exceed traditional approaches [2, 3, 4, 9]. However, evidence is fragmented: datasets, validation schemes, and metrics differ; many studies use small or internally validated cohorts that risk overfitting; and reporting is inconsistent [10, 11, 2, 9].

To provide a clearer picture, we conducted a systematic review and meta-analysis guided by PRISMA - [12]. Our primary outcome is the area under the ROC curve (AUC-ROC), with sensitivity, specificity, precision, and F1 as secondary measures. We tested three hypotheses: DL outperforms classical ML in AUC-ROC [2, 3, 4]; multimodal inputs (e.g., MRI, PET, CSF, clinical) outperform unimodal approaches [1, 13, 8]; and (iii) externally validated studies report lower, more realistic performance than internal-only evaluations [11, 9]. Through pooled estimates and subgroup analyses, we benchmark current performance and highlight the practices needed for clinical translation [2, 8, 14].

## 2 Methods

### 2.1 Plain-Language Summary of Methods

In simple terms, this study combined the results of many individual research papers that used artificial intelligence to detect Alzheimer’s disease. Each study reported how accurately its model could distinguish between healthy individuals, patients with mild cognitive impairment, and those diagnosed with Alzheimer’s disease. We collected these results, along with other key characteristics such as type of input used (MRI, PET, Combination, etc), dataset, sample size and other performance metrics. Using a meta-analytic approach, we then calculated an overall average of model performance while accounting for variation between studies and possible biases and limitations of this method. This approach helps identify consistent trends across the research field, rather than relying on the results of any single study.

### 2.2 Protocol and Reporting

This systematic review and meta-analysis was conducted and reported in accordance with the PRISMA 2020 statement[12] shown in Fig. 1. The review protocol, inclusion criteria, and analysis plan were pre-specified and recorded on the Open Science Framework (OSF) prior to data extraction. All steps from literature search to synthesis were performed in accordance with reproducible research practices and are documented in the public project repository.

**Figure 1:**
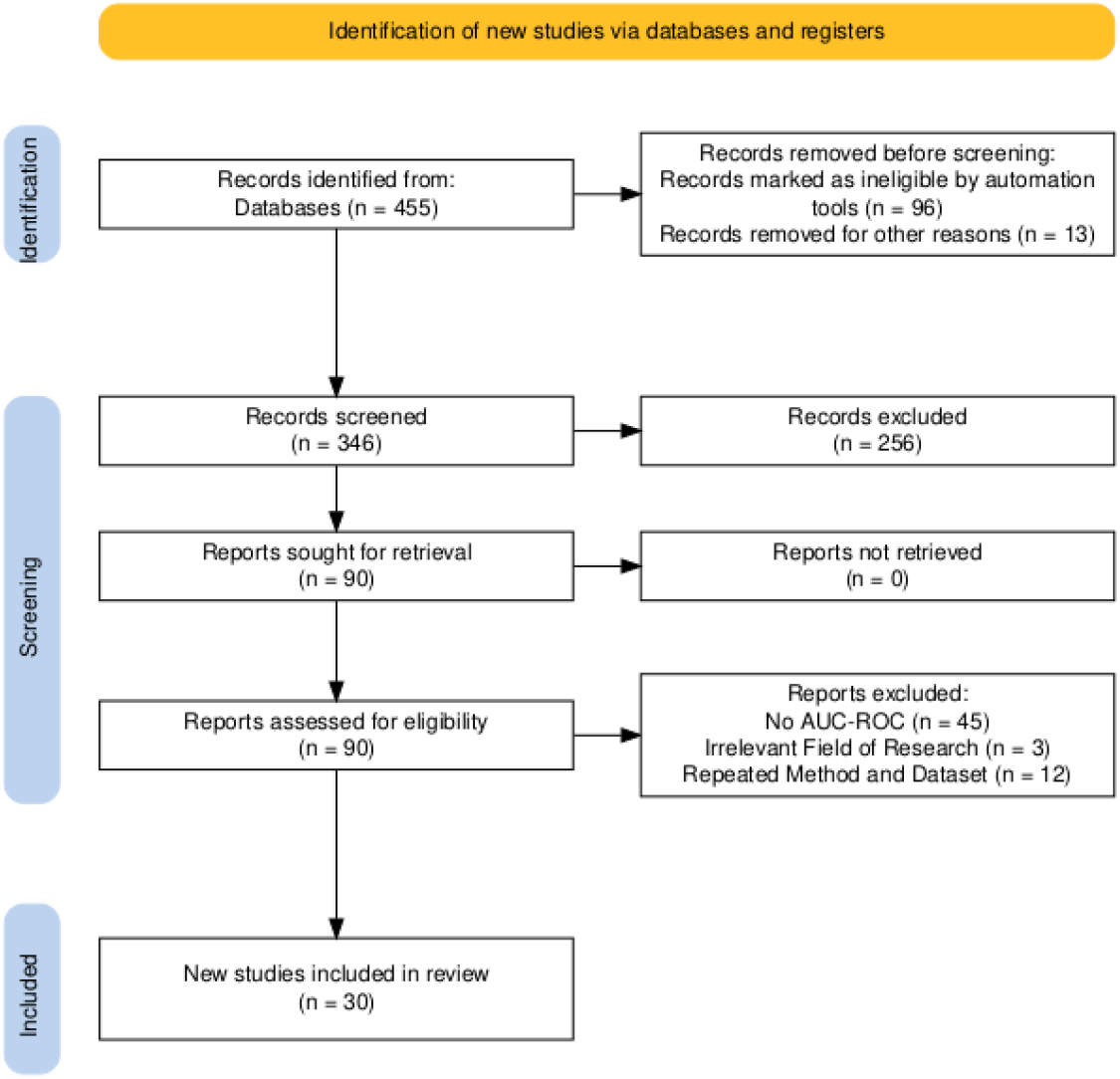
PRISMA 2020 flow diagram illustrating the study selection process. The database search initially identified all retrieved records. After removing duplicates and applying eligibility criteria, 30 studies were included in the final quantitative synthesis (meta-analysis).

### 2.3 Eligibility Criteria

Studies were included if they: (i) used human data; (ii) implemented ML or DL methods for the diagnosis or prognosis of AD/MCI from CN participants; (iii) reported AUC-ROC with optional Sensitivity, Specificity, and F1; (iv) were published in english; (v) published within the past 10 years Exclusion criteria: non-original articles (reviews, editorials), non-diagnostic or non-human studies, conference abstracts without full text, or studies without sufficient quantitative metrics for synthesis. [Table 1] We excluded six full-text articles at eligibility because they were non-original reviews/scoping analyses, methodological critiques, or did not report diagnostic outcomes for AD/MCI vs CN; details appear in Supplementary S5.

**Table 1:**
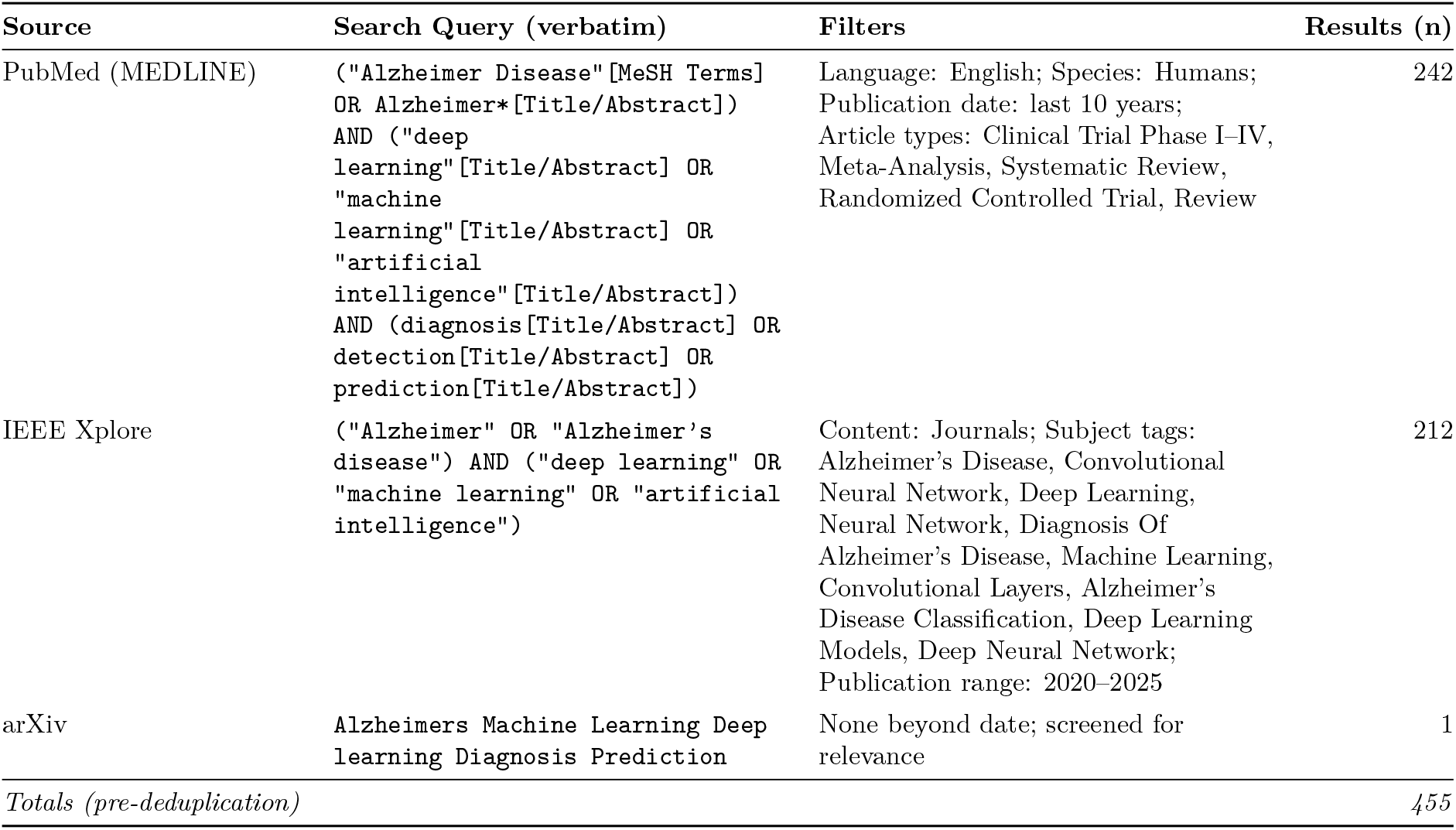
Database search implementations and yields (final search date: 2025-11-10).

| Source | Search Query (verbatim) | Filters | Results (n) |
| --- | --- | --- | --- |
| PubMed (MEDLINE) | ("Alzheimer Disease"[MeSH Terms]<br>OR Alzheimer*[Title/Abstract])<br>AND ("deep<br>learning"[Title/Abstract] OR<br>"machine<br>learning"[Title/Abstract] OR<br>"artificial<br>intelligence"[Title/Abstract])<br>AND (diagnosis[Title/Abstract] OR<br>detection[Title/Abstract] OR<br>prediction[Title/Abstract]) | Language: English; Species: Humans;<br>Publication date: last 10 years;<br>Article types: Clinical Trial Phase I-IV,<br>Meta-Analysis, Systematic Review,<br>Randomized Controlled Trial, Review | 242 |
| IEEE Xplore | ("Alzheimer" OR "Alzheimer's<br>disease") AND ("deep learning" OR<br>"machine learning" OR "artificial<br>intelligence") | Content: Journals; Subject tags:<br>Alzheimer's Disease, Convolutional<br>Neural Network, Deep Learning,<br>Neural Network, Diagnosis Of<br>Alzheimer's Disease, Machine Learning,<br>Convolutional Layers, Alzheimer's<br>Disease Classification, Deep Learning<br>Models, Deep Neural Network;<br>Publication range: 2020–2025 | 212 |
| arXiv | Alzheimers Machine Learning Deep<br>learning Diagnosis Prediction | None beyond date; screened for<br>relevance | 1 |
| *Totals (pre-deduplication)* |  |  | 455 |

**Table 2:**
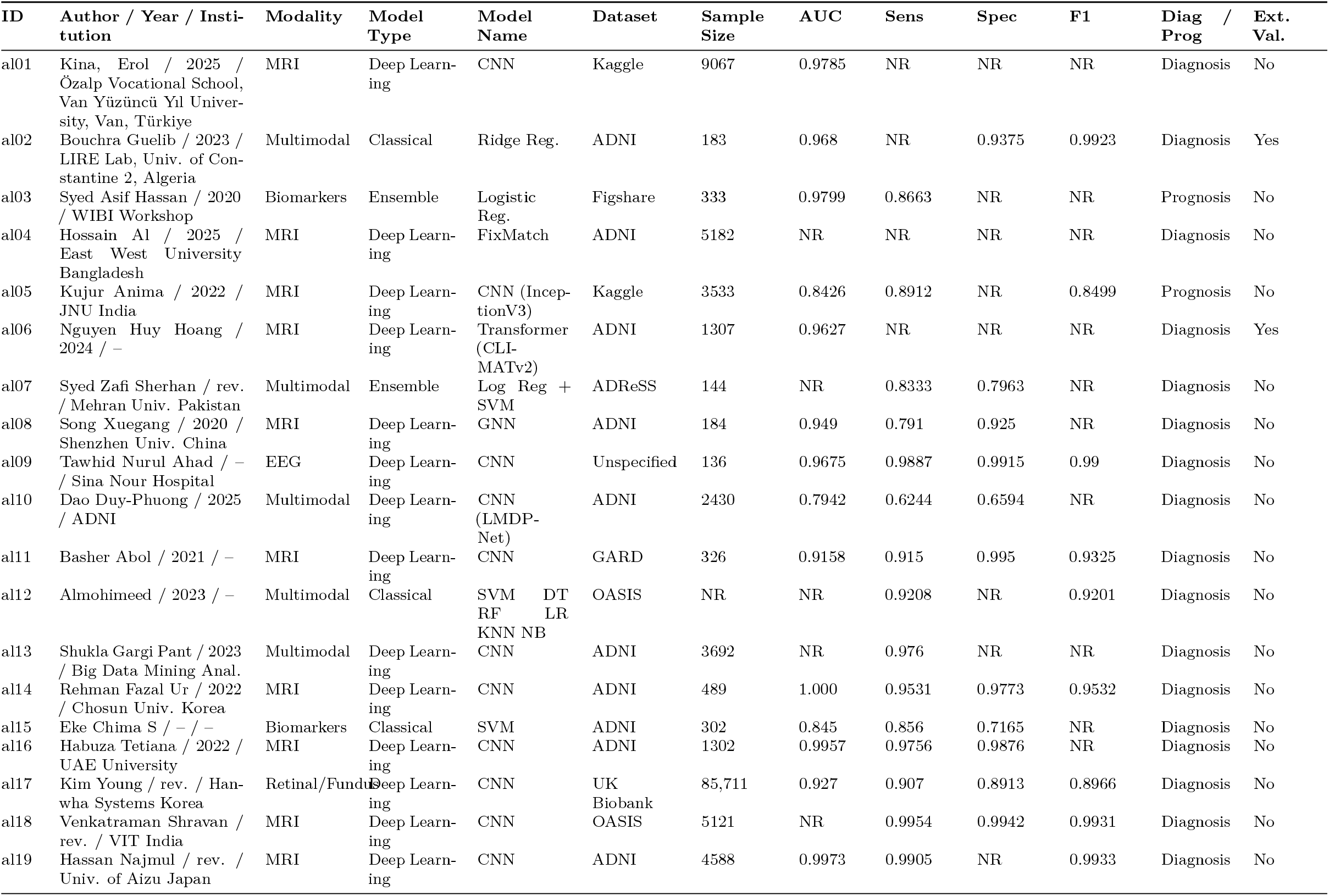
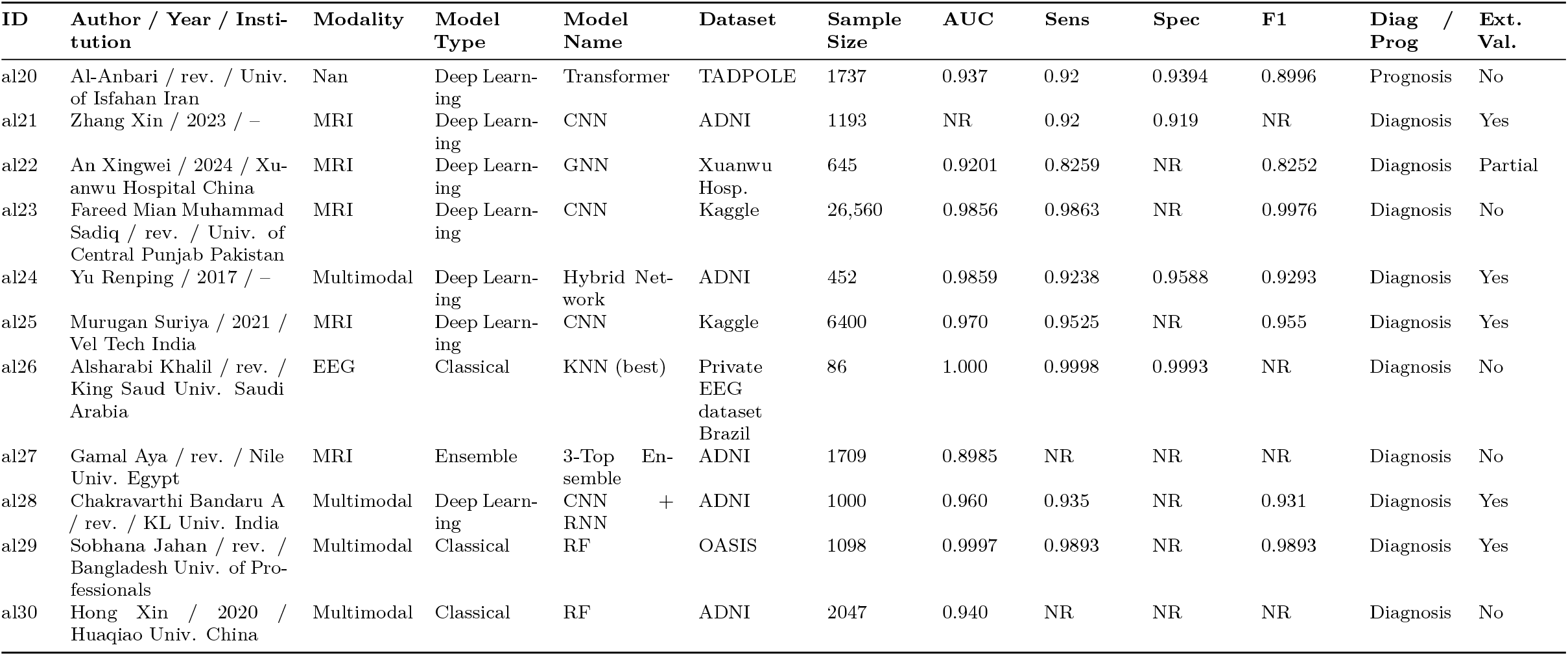
Summary of included studies with key characteristics and reported metrics. “NR” denotes not reported.

### 2.4 Information Sources and Search Strategy

The searches were performed in PubMed (MEDLINE), IEEE Xplore, and arXiv. Keywords combined disease-related terms (“Alzheimer*”, “MCI”), methodological terms (“machine learning”, “deep learning”, “neural network”, “transformer”), and performance terms (“AUC”, “sensitivity”, “specificity”, “diagnosis”). The search window spanned January 1, 2015, to the most recent query date. The references of relevant reviews were manually screened to capture additional eligible studies. [Table 1]

### 2.5 Selection Process

All retrieved records were imported into Rayyan for deduplication and independent screening. Titles and abstracts were screened in the first phase; full-text eligibility was assessed in the second phase using the pre-specified inclusion and exclusion criteria. From which a final text-screening resulted in thirty studies meeting the final inclusion criteria, 25 real AUC contained studies and 5 qualitative discussion studies (See Supplement S2).

### 2.6 Data Collection and Extraction

A standardized data-extraction template was developed and piloted prior to use. Extracted fields included: study ID, author/year/institution, imaging modality, dataset name(s), total sample size, class distribution, data split (train/validation/test), use of external validation, preprocessing and augmentation methods, model family and architecture, pretraining, explainability method, hyperparameter optimization strategy, and reported metrics (AUC, sensitivity, specificity, F1, precision). [2] Where available, confusion matrices were extracted to calculate missing metrics. Missing numeric fields were coded as “NR” and handled as missing values.

### 2.7 Effect Measures and Variance Estimation

The primary effect measure was the study-level area under the ROC curve (AUC). The AUC were transformed to the logit scale due to the clustering of AUC metrics at high values (> 0.9) and thus making meaningful data analyzes difficult. Listed below is the method we used to exemplify the smaller changes in AUC’s:

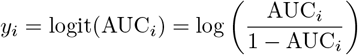

Sampling variances were approximated using the delta method:

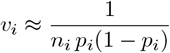

where *n*_*i*_ denotes the effective sample size and *p*_*i*_ = AUC_*i*_. In si

Subgroup analyses compared the pooled AUCs across (a) model families (classical ML vs DL), (b) imaging modalities (MRI, PET, EEG, multimodal), and (c) validation strategy (internal vs external). Analysis was then performed to determine the effects that publication year, sample size, validation and different datasets had across the studies.

Diagnostic performance across studies was primarily expressed using the area under the receiver operating characteristic curve (AUC-ROC), which quantifies a model’s ability to discriminate between classes across all possible classification thresholds. An AUC of 0.5 corresponds to random guessing, whereas an AUC of 1.0 indicates perfect classification.

In addition to the AUC, individual studies frequently reported other measures derived from a confusion-matrix. In essence quantifying a models performance in accurately diagnosing AD/MCI along with being able to set apart the control group for CN patients. They are calculated in the following methods:

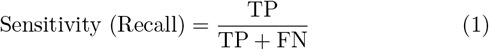

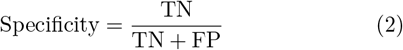

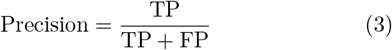

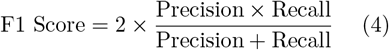

Where TP reflects number of correctly classifying an AD/MCI patients, FN; the number of incorrectly classified CN patients and TN; the number of correctly classified CN patients

Sensitivity reflects a model’s ability to identify positive cases, which in this case consist of identifying AD/MCI accurately. Inversely Specificity is the measure that a model is able to identify a control, a healthy person, from the rest of the population. The final metric F1 represents a harmonic mean between the two values to be a more general indicator of performance.

Together, these complementary metrics provide a multi-dimensional assessment of diagnostic performance: AUC quantifies global discriminative power, while sensitivity, specificity, precision, and F1 score describe performance at a specific decision boundary relevant to clinical application.

### 2.8 Model and Estimation Details

Random-effects pooling used restricted maximum likelihood (REML). AUCs were meta-analyzed on the logit scale; standard errors were approximated via a delta method on AUC (acknowledging that exact variance estimators such as Hanley–McNeil or DeLong require rank-based quantities seldom reported in AI studies). Meta-regression used REML with Knapp–Hartung small-sample correction.

### 2.9 Publication Bias and Small-Study Effects

Publication bias was examined visually with funnel plots of the AUC-ROC performance metric versus its standard error, and quantitatively using Egger’s regression test. Visual inspection of the funnel plot seen in Fig. 2; indicate a distinct asymmetry within the publications. This is supported by the Egger’s regression test, which indicates asymmetry with a p = 0.0003 and intercept of 2.48. Exemplifying the potential publication bias across all the studies.

**Figure 2:**
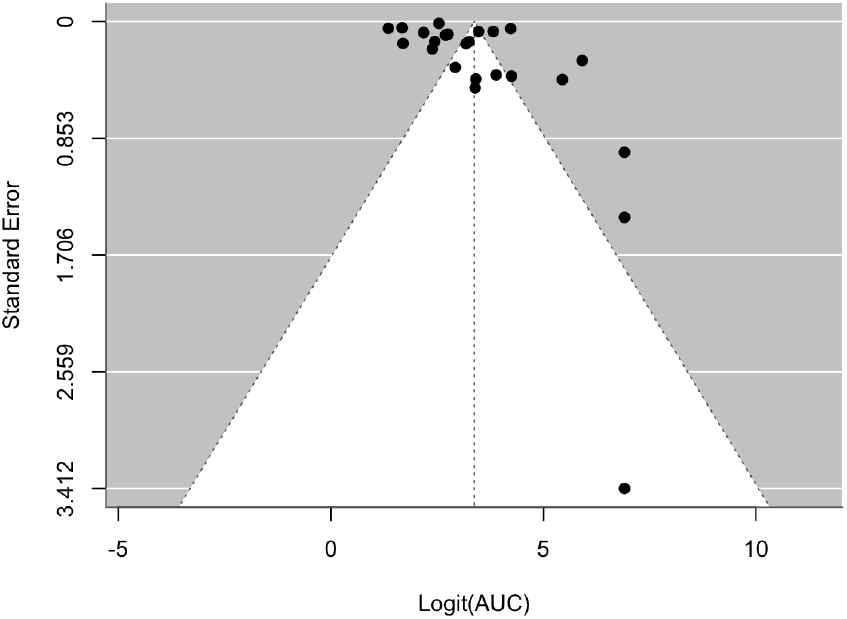
Funnel plot of the logit(AUC) versus standard error with Egger’s test line.

### 2.10 Graphical and Exploratory Analyses

Additional figures summarized relationships among study features. Scatterplots of AUC versus sample size (log-scale) visualized small-sample effects. Heatmaps of mean AUCs by model family and modality highlighted multimodal advantages. The correlation matrices showed the relationships between AUC, sensitivity, specificity, and F1-score in all studies.

### 2.11 Software and Reproducibility

All analyses were performed in R (version 4.3.0) [15]. Core packages included metafor [16] for meta-analysis, tidyverse [17] (including ggplot2, readr, dplyr) for data cleaning and visualization, GGally [18] for correlation matrices, and viridis [19] for color scales. Scripts, processed data files, and generated figures are publicly available on the OSF and project GitHub repository.

## 3 Results

### 3.1 Study Characteristics

A total of thirty studies published between 2020 and 2025 met the inclusion criteria for this meta-analysis Fig. 1. Most studies utilized MRIs (sMRI, fMRI, MRI) as their primary imaging modality, reflecting its established role in assessing brain atrophy patterns in AD and MCI. multimodal pipelines that combined MRI with positron emission tomography (PET), cerebrospinal fluid (CSF) biomarkers, or neuro-psychological test scores formed the second largest group. Smaller subsets employed PET-only, electroencephalography (EEG), or retinal fundus imaging to detect disease-associated alterations.

Deep learning (DL) models were by far the most popular, accounting for over half of all included studies. Convolutional neural networks (CNNs) were the most frequently adopted architecture, followed by transformers, graph neural networks (GNNs), and ensemble methods. A smaller portion of papers used traditional machine learning algorithms such as support vector machines (SVMs), random forests, and logistic regression.

Datasets were overwhelmingly derived from large, publicly available repositories such as the Alzheimer’s Disease Neuroimaging Initiative (ADNI) and the Open Access Series of Imaging Studies (OASIS), with several institution-specific EEG and PET datasets supplementing smaller cohorts. Sample sizes ranged widely: from fewer than 100 to more than 85,000 participants, with most studies focusing on differentiating AD, MCI, and cognitively normal (CN) groups.

External validation, defined as model testing on independent datasets not used during training or parameter tuning, was reported in fewer than one-third of included studies. The majority relied solely on internal crossvalidation or hold-out test sets. This imbalance suggests that while reported accuracies are high, generalization across unseen cohorts may remain limited.

### 3.2 Overall Diagnostic Performance

Random-effects pooling on the logit scale produced a high overall discriminative accuracy, yielding a back-transformed pooled AUC of 0.962 (95% CI: 0.939–0.977) [Full formulas in Supplementary Section S1]. Individual study estimates and the pooled random-effects mean are summarized in Fig. 3. Confidence intervals were generally tighter for MRI and multimodal studies, while PET and EEG-based models displayed broader ranges, reflecting smaller sample sizes and higher methodological variability.

**Figure 3:**
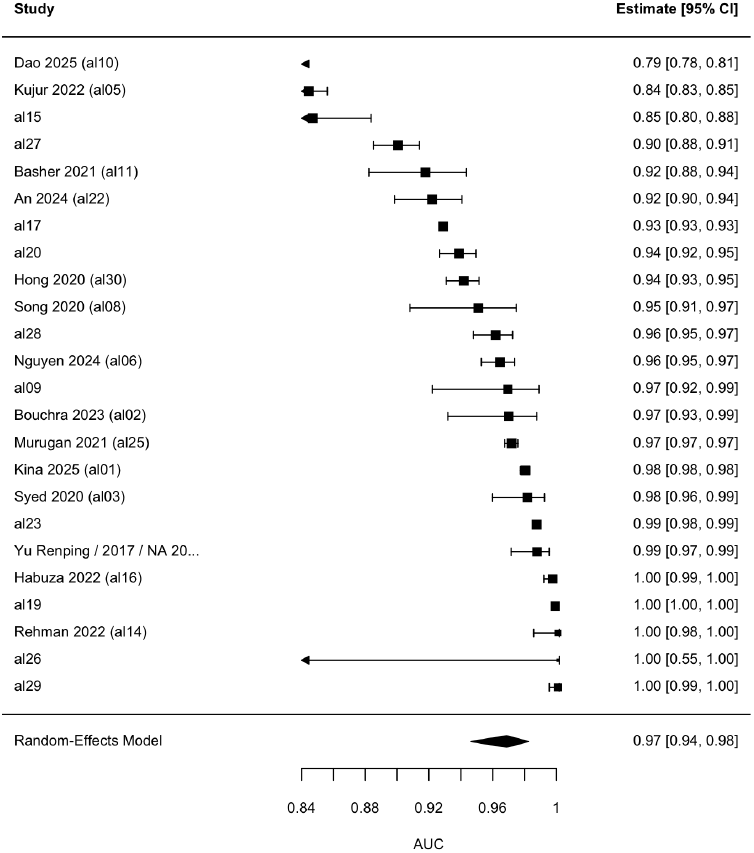
Random-effects forest plot of study-level AUCs. Error bars represent 95% confidence intervals; the diamond indicates the pooled estimate.

Across studies reporting operating characteristics, the mean sensitivity was 0.914, mean specificity was 0.913, and the average F1-score was 0.94. These results indicate balanced performance between true positive and true negative detection rates, suggesting that most models were successful in predicting both AD/MCI patients about as well as CN patients. These metrics confirm that DL and ML classifiers are very capable of accurately distinguishing each type of patient in their respective controlled datasets.

### 3.3 Subgroup Analyses

#### Model family

Performance remained uniformly high across model families as shown in Fig. 4. Ensemble methods achieved slightly lower median AUCs (*≈* 0.93) compared to DL and classical ML models, which reached median AUCs of *≈* 0.96 and *≈* 0.97, respectively. However, the substantial overlap across their distributions suggests that architectural differences between models play a less important role relative to other factors such as sample size, validation testing rigor, if the data was pre-processed or not, and the different types of imaging modalities present throughout the study.

**Figure 4:**
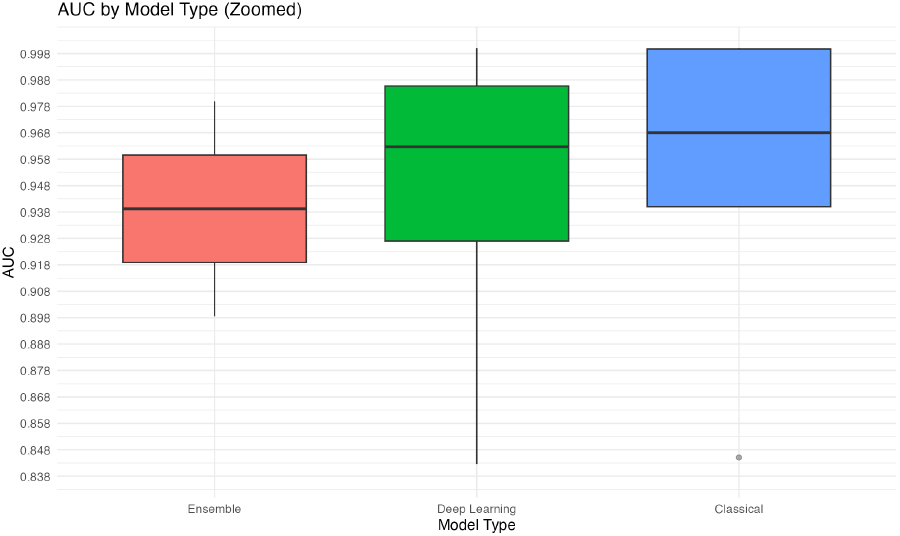
Distribution of AUCs by model family. Medians overlap substantially, indicating limited performance separation among architectures.

### 3.4 Imaging modality

AUC distributions were high across all imaging modalities, Fig. 5. Models that were trained on EEG data achieved the highest median pooled AUC (*≈* 0.97), closely followed by multimodal and MRI models (*≈* 0.965). The elevated performance in these 3 categories highlights the effect of training models on complex structural and physiological information (EEG’s and MRI’s) or a combination of both to enhance accuracy.

**Figure 5:**
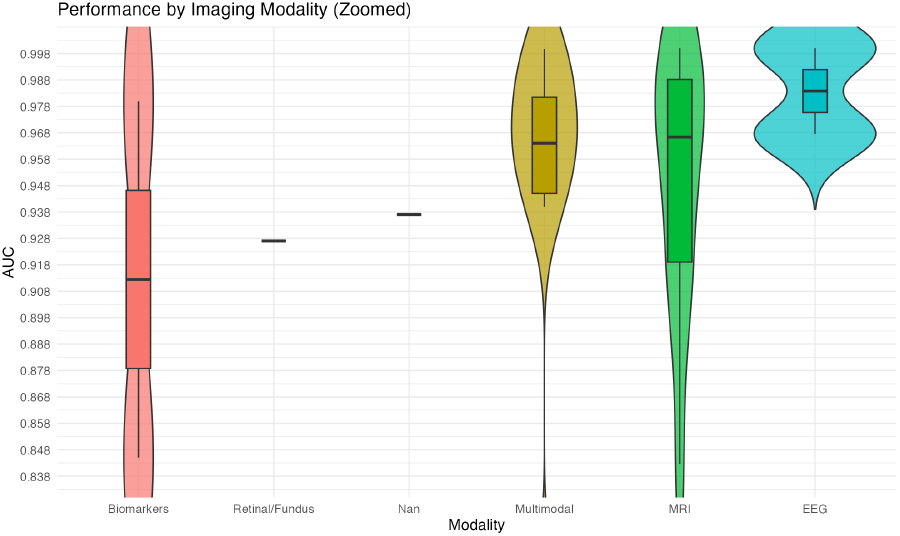
AUC distribution by imaging modality. multimodal pipelines show a mild advantage but overlap with MRI-only workflows.

**Figure 6:**
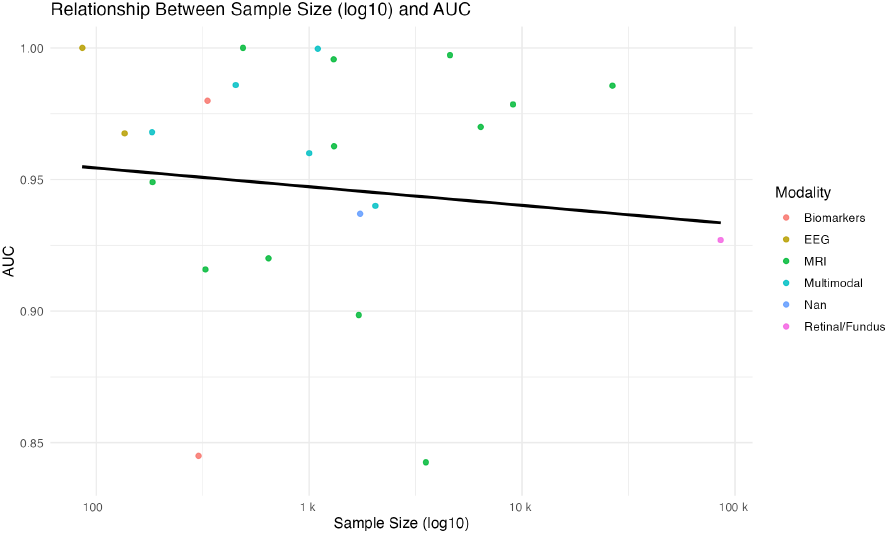
Scatter plot of sample size (log_10_ scale) versus AUC. High accuracy is observed across dataset sizes, indicating possible performance inflation in small-sample studies.

### 3.5 Sample size and reported accuracy

Although not strong, a negative trend in the median AUC was observed as sample size increased; Fig. 5. No-tably, the 95th percentile AUC dropped significantly for studies with smaller samples, a phenomenon that typically indicates inflated validation estimates or overfitting due to limited datasets. Conversely, the larger samples demonstrated conservative but far more credible AUCs, with the median performance stabilizing at approximately 0.94. The consistent and high median AUC across the entire sample size range further implies that, despite the risk of small-sample inflation, models are generally effective at both small-sample and large analysis scales.

### 3.6 Model–modality interactions

The heatmap Fig. 7 summarizes mean AUCs across model and modality pairings. While a Deep Learning approach applied to MRI data was the most common pairing (*n* = 11) with a strong mean AUC of 0.956, the highest mean AUCs were observed for Classical models with EEG (1.000) (Note that the EEG model that achieved this was [20]; which used a private and comparatively small dataset) and Ensemble models with Biomarkers (0.980), though these were based on single studies. The multimodal-Classical pairing also demonstrated a high mean AUC of 0.969 (*n* = 3). The marginal difference between performance across different model-modality pairings emphasize that current technology related to diagnosis of AD/MCI has reached saturation across many different types of both data and architecture.

**Figure 7:**
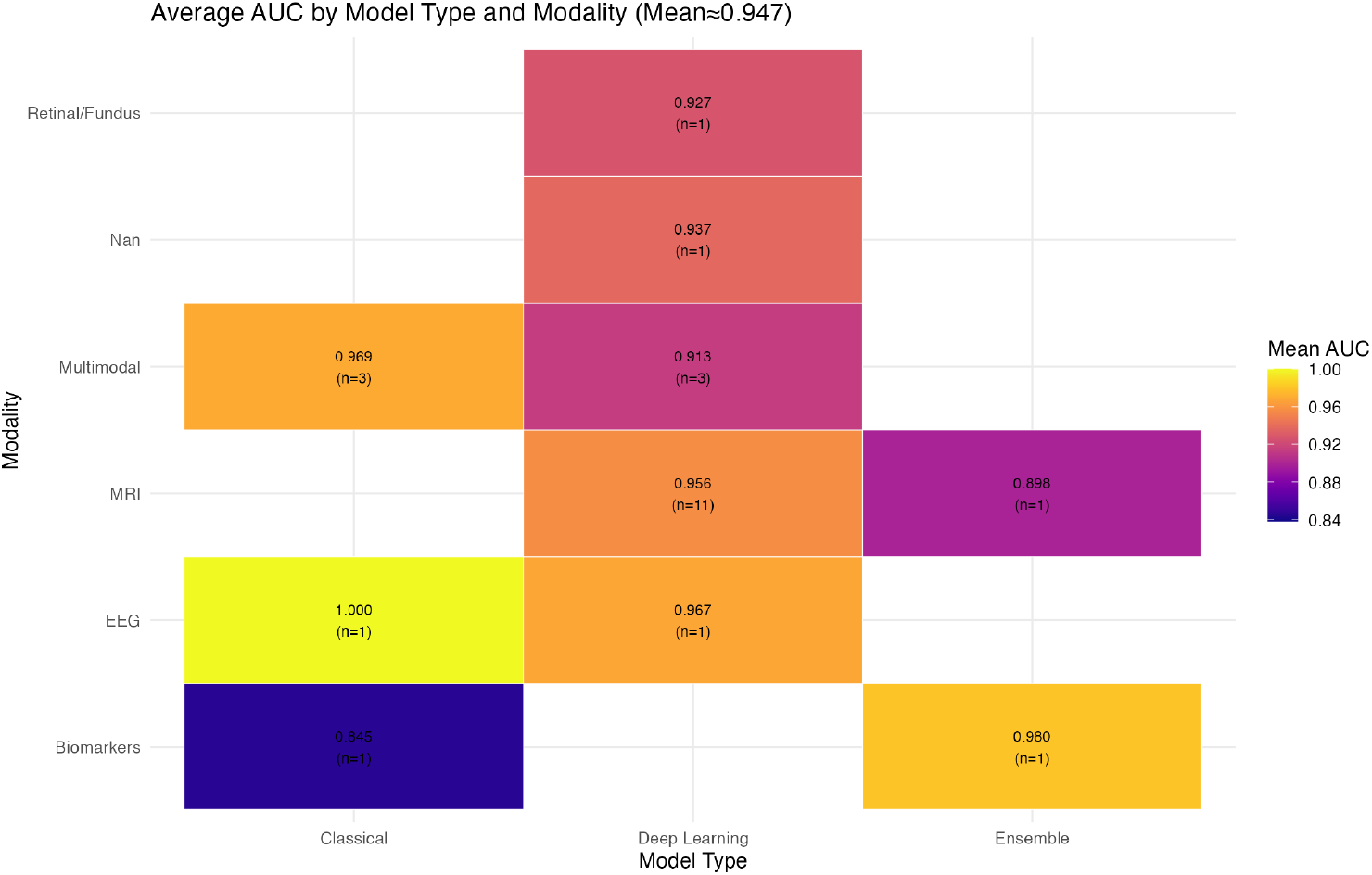
Mean AUCs by model family and imaging modality. CNNs paired with MRI and multimodal inputs dominate the top-performing cells, though differences are minor overall.

### 3.7 Temporal performance trends

Temporal analysis of the AUC over time shows us a non-monotonic progression of AUC, challenging the expectation of increasing performance over time. Reported AUCs declined from 0.98 in pre-2018 studies to 0.89 in several post-2023 reports Fig.. 8. The observed decrease in AUC and performance can be described by the growing focus on generalized multimodal algorithms such as Dao et al. [13], which uses 5 different input formats in its model. Indicating a shift toward more robust, clinically representative benchmarks that are less prone to optimistic bias from highly curated, narrow-scope studies.

**Figure 8:**
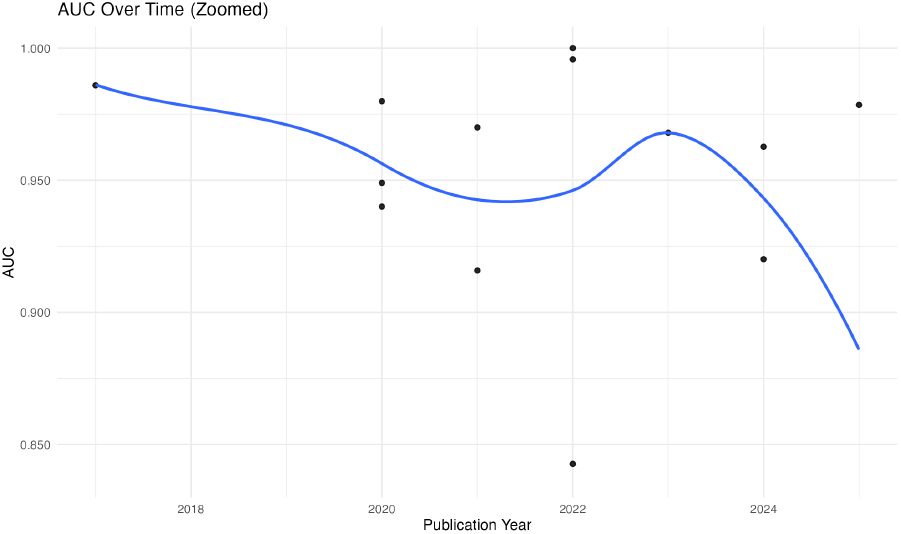
Temporal trend of reported AUCs from 2020–2025. Performance rose through 2023 and stabilized near a ceiling, indicating maturation of the field.

### 3.8 Operating characteristics and metric correlations

The relationship between sensitivity and specificity across studies is displayed in Fig. 9. Most points cluster within the upper right quadrant, confirming the balanced model performance we described earlier. Studies employing external validation tended to yield slightly lower but more conservative estimates, suggesting better generalization across unseen data. This finding aligns with our previous critcism of AUC inflation, a byproduct of smaller datasets with no external validaton.

**Figure 9:**
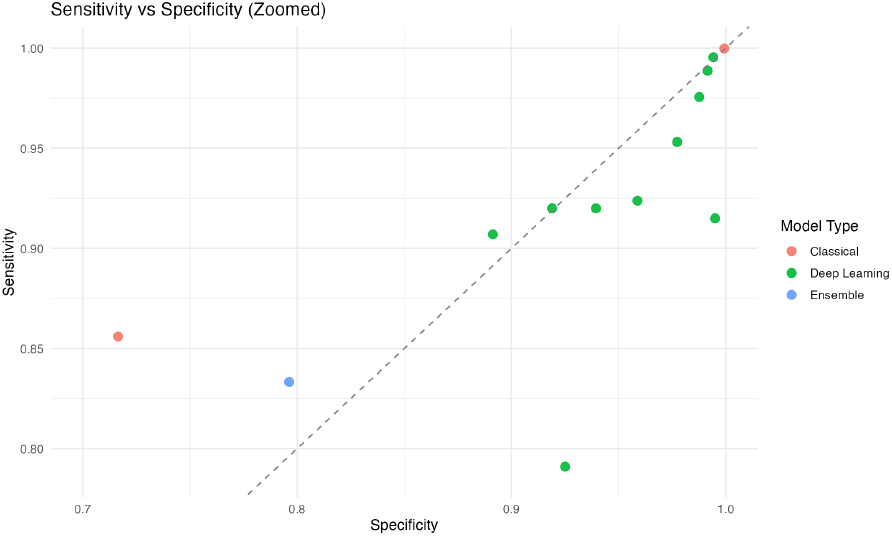
Sensitivity vs. specificity across studies. External validation cases trend lower but remain more consistent across cohorts.

The correlation analysis among key discriminative and operational metrics such as AUC, Sensitivity, Specificity, and F1-score Fig. 10,revealed strong to extremely strong positive relationships across all pairings. Specifically, Specificity showed the highest correlation with AUC (Corr: 0.918), followed by F1-score (Corr: 0.781) and Sensitivity (Corr: 0.772). The high positive correlations, especially the strong relationship between AUC and F1-score, suggest that the robust discriminative ability reported in high-AUC studies translated into balanced operational behavior across classes. Furthermore, the very strong correlation between Sensitivity and Specificity (Corr: 0.838) indicates that high performance on one class was generally not achieved at the expense of the other, reinforcing the overall reliability and robustness of the top-performing models.

**Figure 10:**
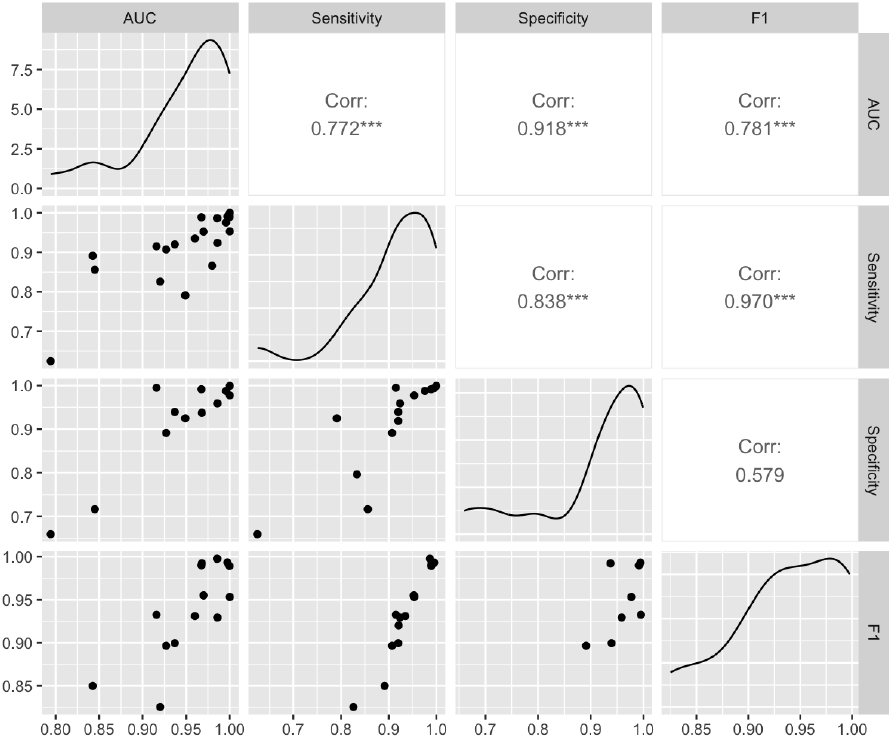
Correlation matrix among AUC, sensitivity, specificity, and F1-score. Strong positive correlations reflect consistent multi-metric performance.

### 3.9 Summary of Robustness and Key Findings

Overall, the synthesis of thirty recent studies, informed by quantitative analysis of performance metrics, demonstrates three major trends:

1. **Performance Saturation with Methodological Nuance**. While reported AUCs demonstrate uniformly high discriminative performance across modalities (median *≈* 0.94), this masks significant volatility. The strong positive correlations among AUC, Specificity, and F1-score confirm that high discriminative ability translates to balanced operational performance, collectively signaling maturity and saturation of models and architecture to predict AD and MCI.
2. **Validation Rigor and Data Independence**. A crucial observation from the relationship between sample size and AUC is that high AUCs in small datasets (log_10_ Sample Size *<* 3.5) were often inflated, peaking near 1.00. In contrast, studies incorporating larger, more robust samples demonstrated conservative yet more credible results (median AUC *≈* 0.94). This reinforces that methodological transparency, strict validation protocols, and data independence are critical determinants of reproducibility, more so than architectural and modal differences between specific models.
3. **Modality-Specific Excellence and Multimodal Utility**. The EEG modality demonstrated the highest pooled median AUC (*≈* 0.978), though Multi-modal and MRI-only workflows performed comparably (median *≈* 0.965). Furthermore, the Deep Learning+MRI pairing emerged as the most widely studied and validated pairing (AUC 0.956). This evidence suggests that highly optimized, modality-specific pipelines remain exceedingly viable and effective when multimodal integration is not feasible, often showing little practical difference in peak AUC compared to combined approaches.

Collectively, these results highlight that while technical progress has driven substantial gains in Alzheimer’s disease classification accuracy, the recent sharp post-2023 decline in aggregate AUC suggests a critical shift in research focus. The next frontier will likely depend on tackling more challenging, clinically relevant prediction tasks (prognosis, multiclass differentiation), along with an increased emphasis on harmonized datasets, external benchmarking, explainability, and clinical translation rather than continued raw AUC optimization.

## 4 Discussion

### 4.1 Principal Findings

In this quantitative meta-analysis of 30 different studies taken between 2015 - 2025, we’ve consistently observed high diagnostic performance across modalities and models in differentiating AD and MCI from CN patients. Quantitatively, this is proven as the pooled AUC shows us a model performance score of 0.962. Although EEG is defined as the best performing modality with a median AUC of 0.978, it should be noted that only 2 of the 30 studies were EEG-based [5] [20], of which both had extremely low sample sizes of 136 and 86, respectively. Also, noting that both datasets were privately obtained, therefore questioning the real credibility of EEG diagnosis methods. MRI and Multimodal methods, however, are well documented and consistently show high AUC, showing consistency and maturity of image-based classification approaches to diagnosing AD/MCI. Even the discrepancy between the AUC of smaller studies and larger studies is low, suggesting models can reach exceptional AUC irrelevant of the dataset. However, taking a look at Fig. 6 we can see that lower sample sizes can lead to inflated AUC, potentially indicating instability and overfitting.

Although a majority of studies reported exceptionally high AUC, the zoomed-scaled visualizations allowed us to find subtle, yet significant, differences across both model architectures and modalities. The Deep Learning and MRI pairing emerged as the most robust and validated approach [21] [22], posting a mean AUC of 0.956 across 11 different models. However, the magnitude of performance between major models and modality pairings was minimal, indicating that the discriminative ability has/or is reaching saturation in terms of AUC. Leading us to believe that other factors, such as datasets, pre-processing, and methodology, may drive more variance than model architecture.

Our analysis of other characteristics other than AUC to determine overall model performance and to gain diversity of success indicates that AUC, Sensitivity(Recall), Specificity, and F1-Score have a strong positive relationship. Suggesting that gains in discriminative performance were generally not preceded by trade-offs in other discriminative qualities. One such study is Kim et. al [7] whose model had sens of 0.907 and spec of 0.8913 on a dataset of nearly 86,000 people. Emphasizing that the model was able to identify TP (True Positives) just as well as TN (True Negatives) over a vast dataset.

The AUC over time graph Fig. 8 shows us that, contrary to popular belief, the AUC metrics have been steadily declining over time. However, this can be attributed to a shift towards more robust and practical applications of models such as Dao. et al [13], a multimodal CNN-based model that takes 5 different types of inputs (MRI, PET. Biomarkers, Genetic, and Demographic) all unlabeled, to diagnose MCI/AD. A novel method that indicates a shift from hyper-specific database-reliant and self-validating models to more practical and broad usecases for potential clinical and general use.

### 4.2 Comparison with prior work

This systematic review and meta-analysis builds off of the work of other previous analyses by broadening the scope and focusing on one singular metric, the AUC-ROC, over accuracy. Allowing for a more comprehensive overview of the field than mixing different factors over subfields. For example in Ozkan et al. [23] they discuss the same lack of datasets and say that DL shows “promise … further research is needed” for longitudinal cross-population validity. The meta-analysis conducted in this study excluded the specification of subgroups and pooling over datasets. Another similar meta-analysis [24] also describes inconsistency in data collection and worked with traditional OCT/OCTA images. However, choosing to focus on the industry as a whole we eliminate the specificity in these two analyses and are able to evaluate the industry quantitatively as opposed to qualitatively like these two meta-analyses.

### 4.3 Quality, Bias and Generalizability

Our analysis indicates several quality and bias concerns. Firstly the lack of widespread external validation remains a significant concern, with data implying that it may systematically over-report performance metrics. The second concern is the credibility associated with the lack of large datasets. Models trained on smaller datasets have been shown to inflate AUC as a result, this is particularly shown in models that use some kind of pre-processing such as SMOTE and SSMI on their smaller dataset. Furthermore another concern is the general homogeneity of data sets that are observed. ADNI and OASIS datasets make up the vast majority of datasets that models are trained upon. Limiting the generalization of such models and removing transiency to a realistic clinical population with more ambiguous and non-perfect data. Lastly the concern of success-bias reporting as the Egger Test determined that there was bias, we can see that since most of the AUC’s for models written exist in the upper echelon of 1 - 0.9, an indication of publication bias. For more information see Supplementary S1.8

Despite all of this, our analysis suggest that elevated AUC generally corresponds to elevated sens, spec, and F1, suggesting that class imbalance or bias towards a particular outcome isn’t widespread. Nevertheless, reproducibility is a primary concern for models without external validation and the more conservative metrics from externally validated studies [25] are more stable and credible estimates, while also posting a >0.95 AUC.

### 4.4 Implications and future directions

The findings of this synthesis do prove our Hypothesis that ML and DL models have a >0.8 AUC when it comes to diagnosing AD/MCI, however the lack of diversity in datasets along with the AUC inflation of smaller and non-externally validated models raises concerns to the validity of the mean 0.94 AUC we found. For the future there must be stricter guidelines on data, inter-model comparison, dataset diversity and other benchmarking protocols to ensure increased reproducibility. This mandate includes harmonized image registration, intensity normalization, and feature extraction strategies. Second, prospective multi-center validation studies are urgently needed to confirm model generalizability and to identify potential cohort-specific performance differences that would impede real-world adoption. Third, future research should transition away from simple cross-sectional diagnostic labels and incorporate clinically grounded endpoints, such as predicting conversion from MCI to AD or conversion to dementia over longitudinal follow-up. Fourth, reporting standards must be rigorously enhanced to mandate the inclusion of complete train/test splits, confusion matrices, confidence intervals, and external validation outcomes. Fifth, Explainable AI (XAI) approaches and visualization tools are paramount to facilitating clinical adoption and trust, enabling physicians to understand the model’s rationale. The integration of multimodal data, rigorous validation, and transparent reporting will be critical for clinical translation. Although discriminative performance is approaching a ceiling in current studies, the next wave of AI research must prioritize interpretability, clinical utility, and scalability over marginal AUC gains. Future studies should strategically explore advanced models, such as hybrid architectures combining graph-based reasoning with convolutional feature extraction, or transformer-based temporal modeling, to capture the dynamic progression of AD pathology beyond static classification tasks. Ultimately, AI tools for AD must be designed not only for accuracy but also for robustness, fairness across diverse demographic subgroups, and ease of integration into existing clinical workflows.

## 5 Limitations

Several limitations of this meta-analysis need to be considered during the final product. While the gold-standard approach for diagnostic accuracy synthesis is a bivariate model on sensitivity and specificity, most AI-based Alzheimer’s studies report only AUCs without full confusion matrices. Thus, we used a random-effects model on reported AUCs to summarize performance trends, acknowledging that this simplifies threshold effects and may overestimate generalization. Secondly, reporting heterogeneity across studies complicates pooled estimates as not all studies provided per-class counts, SD, confidence intervals or other such metrics. Of which we required approximation and recalculation of those specific metrics. Thirdly, the limited access to the train-test-validate splits restricted the ability to fully harmonize pre-processing and evaluation procedures. Another glaring limitation is that of the EEG and PET modalities, which only consist of very few studies out of the majority, therefore artificially inflating their AUC and posing a disproportionate influence if over-represented in pooled analyses. Lastly we discussed the publication bias that is present due to Egger’s test, showing that an AUC >0.9 was the most common, suggesting that DL and ML architectures are more likely to be accepted if performance metrics were favorable. Despite these limitations, the inclusion of thirty studies with diverse modalities, architectures, and validation strategies provides a comprehensive snapshot of current AI-based diagnostic performance, and the results remain informative for guiding future work.

## 6 Conclusion

### 6.1 Key Findings and Implications

- Performance Saturation: AUC analysis confirmed that performance is approaching a ceiling, with the majority of well-validated models clustering around a conservative mean pooled AUC of 0.962. While the DL–MRI pairing proved the most robust (0.956 mean AUC across 11 studies), differences between top-performing methods were modest. Implying the neglible differences between different model architectures and modality pairings.
- Methodological Rigor and Bias: The observation of AUC inflation in small-sample studies and the sharp decline in aggregate AUC post-2023 are significant findings. This decline reflects a necessary shift in research focus toward more complex models which adopt a more generalized multimodal system that is better suited for general clinical applications.
- Balanced Operational Behavior: Strong correlation between AUC, Spec, and F1 indicated that often models that were able to accurately classify patients of AD or MCI, were able to do so with similar accuracy for CN patients.

### 6.2 Future Directions for Clinical Translation

For successful clinical translation, emphasis must shift from simple AUC optimization to methodological rigor and interpretability. Future research must prioritize:

1. Independent External Validation: Required to ensure that high performance is maintained across diverse, real-world patient populations, mitigating the biases introduced by ADNI/OASIS dataset homogeneity.
2. Explainable AI (XAI): XAI strategies and interpretable outputs are paramount to facilitating physician trust and clinical adoption.
3. Reporting Transparency: Standardized pipelines and rigorous metric reporting are essential for ensuring reproducibility and safe clinical implementation.

Overall, this synthesis reinforces the immense potential and success of AI-based diagnostic systems to enhance early detection of AD/MCI, while simultaneously underscoring that the next frontier of progress lies not in marginal gains in AUC, but in methodological robustness, generalizability, and interpretability.

## Data Availability

All data produced in the present work are contained in the manuscript. Extracted study-level data and R analysis scripts are publicly available on the Open Science Framework project repository.

https://osf.io/asy2j/overview

https://github.com/sakisaki-dev/Deep-Learning-for-Early-Detection-of-Alzheimer-s-Disease-Meta-Analysis-

## Abbreviations

AD: Alzheimer’s Disease
MCI: Mild Cognitive Impairment
ML: Machine Learning
DL: Deep Learning
AUC/AUC-ROC: Area Under the Curve Receiver Operating Characteristic
MRI: Magnetic Resonance Imaging
PET: Positron Emission Tomography
EEG: Electroencephalography
CI: Confidence Interval
AI: Artificial Intelligence
CN: Cognitively Normal Patients
SMOTE: Synthetic Minority Oversampling Technique
SSMI: Similarity-based Modality Integration
Sens: Sensitivity
Spec: Specificity
TP: True Positive
TN: True Negative
FN: False Negative
FP: False Positive

## Conflict of Interest and Author Contributions, Ethics and Funding

### Author Contributions

Saketh Machiraju conceived the study design, conducted the systematic search and data extraction, performed the statistical analysis, interpreted the results, and wrote the manuscript. All analyses, figures, and text were completed independently.

### Ethics

Secondary analysis of published, de-identified data; IRB review not required.

### Funding

No external funding was received.

## 7 Supplement

### S1.1 Data harmonization

We read the extraction CSV and normalized fields as in the public code: string cleaning to handle “NR”/”N/A”, normalization of modality (`MRI, PET, EEG, fMRI, Retinal/Fundus, Multimodal`), model type (e.g., `CNN, GNN, Transformer, Ensemble, Classical ML`), and external validation (`Yes/No/Partial`). We parsed numeric `sample_size_total, auc_roc, sensitivity, specificity, f1` and derived the publication year from the author string.

### S1.2 Study inclusion for meta-analysis

For pooling, we kept studies with 0 *<* AUC *<* 1 and non-missing `sample_size_total` *>* 3. If multiple outcomes per paper were available, we used the single AUC provided in the extraction table. All other summaries (e.g., sensitivity/specificity means, subgroup plots) include any records with the corresponding metric available.

### S1.3 Effect size and within-study variance

We transformed AUCs to the logit scale after clipping to avoid infinities:

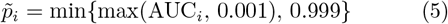

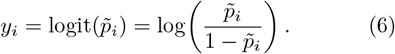

Within-study variances for *logit*(AUC) were approximated for all studies by a delta-method formula using the total sample size *n*_*i*_ as the effective *n*:

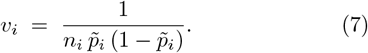

**Note**. Alternative AUC variance estimators (e.g., Hanley–McNeil, DeLong) were *not* used because per-class counts or score distributions were not consistently available in the dataset; the analysis uses the delta-method throughout.

### S1.4 Random-effects model (REML)

We fit a random-effects model on *y*_*i*_ with within-study variance *v*_*i*_:

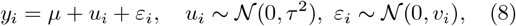

using restricted maximum likelihood (`metafor::rma(yi, vi, method=“REML”)`). We report Cochran’s *Q, τ*^2^, and *I*^2^ = max *{*0, (*Q −* (*k−* 1))*/Q}*. Pooled estimates and confidence intervals from `rma` were back-transformed to the AUC scale with the inverse logit. Knapp–Hartung adjustments and meta-regression were *not* applied.

### S1.5 Small-study effects and funnel plot

We assessed asymmetry on the logit scale using Egger-type regression `via metafor::regtest(res, model=“rma”)`. Let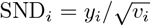 and 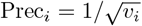 The test regresses

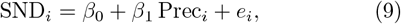

where a nonzero intercept *β*_0_ indicates funnel-plot asymmetry. Funnel plots were drawn on the logit(AUC) axis with standard error on the *y*-axis.

### S1.6 Forest plot and back-transformation

We used `metafor::forest` with `transf = transf.ilogit` to display individual logit(AUC) estimates on the AUC scale alongside the REML pooled estimate. For a compact figure, we also computed approximate study-level 95% CIs as 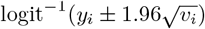 and plotted them with `ggplot2`. Axis limits were “zoomed” by quantile-based bounds to enhance readability of clustered high AUCs.

### S1.7 Descriptive (non-pooled) summaries and graphics

We summarized sensitivity, specificity, and F1 by simple arithmetic means over available studies (no pooling). Subgroup graphics—AUC by model family and by modality—used violin/boxplots of raw AUC values (unweighted). The sample-size relationship used scatter plots with a linear smoother (`geom_smooth(method=“lm”)`) and log_10_ *x*-axis scaling; temporal trends used the default `geom_smooth` (LOESS for these data sizes). The model *×* modality heatmap reports unweighted means with *n* counts. Sensitivity vs. specificity was plotted with zoomed axes. A correlation matrix among AUC, sensitivity, specificity, and F1 was produced with `GGally::ggpairs` using pairwise-complete observations.

### S1.8 Limitations of these choices

(i) Using the total sample size as the effective *n* for AUC can overstate precision under severe class imbalance; results should be interpreted with that caveat. (ii) The delta-method variance treats AUC like a smooth function of an underlying proportion and ignores ranking uncertainty; it is an approximation chosen for completeness across heterogeneous reports. (iii) Subgroup figures and descriptive averages are not meta-analytic estimates and are unweighted by study precision.

### S2 Qualitative Assessment of Non-AUC Studies

Although five studies did not report an explicit AUC-ROC value (al04, al12, al13, al18, al21), their inclusion remains important for understanding the methodological breadth of machine and deep learning research in Alzheimer’s disease (AD) detection. These papers contribute valuable insight into experimental design, modality integration, and model transparency, despite their quantitative exclusion from pooled performance metrics. Hossain et al. focused on a semi-supervised deep learning framework using Fixmatch for MRI-based classification of AD and MCI. While the authors did not provide AUC-ROC scores, they emphasized adaptive pseudolabeling and consistency regularization, demonstrating how unlabeled MRI data can enhance training efficiency and generalization. This highlights an emerging trend in leveraging semi-supervised learning for neuroimaging, where reporting standardized diagnostic metrics remains a challenge [26].

Almohimeed et al. explored a multimodal approach integrating structural MRI and clinical biomarkers using an ensemble of classical machine learning classifiers such as SVM, Decision Trees, and Random Forests. Their study underscored model interpretability and comparative classifier evaluation, providing a useful baseline for multimodal fusion studies even in the absence of AUC reporting. However, the omission of AUC prevents a unified comparison of classifier discriminative ability [31]. Shukla et al. implemented a deep convolutional neural network trained on ADNI multimodal inputs but primarily reported accuracy and sensitivity metrics. The study exemplifies the reliance on accuracy based metrics that, while intuitive, can mask class imbalance effects in AD versus MCI classification tasks. Their contribution lies in illustrating how deep neural feature extraction can improve recall, though standardization with AUC-ROC would aid interpretability [32].

Venkatraman et al. presented an MRI-based CNN architecture emphasizing near-perfect sensitivity and specificity (0.9954 and 0.9942, respectively). Despite lacking an explicit AUC value, the nearly symmetric performance across metrics suggests a potentially high discriminative threshold. The authors’ methodological rigor macro-averaged validation and consistent test splits supports their reliability, but the absence of AUC limits direct comparability with other CNN-based studies [33].

Zhang et al. proposed a deep CNN using ADNI data that focused on reporting sensitivity and specificity without summarizing discriminative power via AUC. Their architecture aligns with contemporary volumetric feature extraction pipelines, but the omission of AUC and precision metrics restricts full assessment of model calibration and balance between positive and negative prediction rates [3].

Collectively, these studies highlight a critical methodological gap: the lack of standardized diagnostic reporting. Their qualitative importance lies in demonstrating that while deep and classical algorithms are converging toward high sensitivity and specificity, consistent use of AUC-ROC (and its derivatives such as partial AUC or precision–recall AUC) remains essential for reliable cross-study comparison. As AI-based diagnostic literature matures, such reporting standardization will be key for reproducibility and integration into meta-analytic frameworks.

### S3 Screening and Extraction Protocol

Screening: Single-reviewer screening in Rayyan; a 20% random sample was rechecked at full text against inclusion rules. Inclusion Rules: Human data; ML/DL diagnosis of AD/MCI vs CN; AUC reported or convertible; English; 2015–2025. Exclusion Rules: Non-diagnostic, animal, abstract-only, insufficient metrics. Extraction Single reviewer, piloted form; “NR” coded as missing; conflicts resolved by pre-specified rules.

### S4 Sensitivity Analyses

Across specifications, pooled AUCs were stable: FE 0.929 vs REML 0.963; excluding AUC *≥* 0.99 gave 0.947; external-only 0.977; raw-scale 0.942. Heterogeneity was high (I^2^ *~* 99%) except external-only (77%); corresponding 95% prediction intervals were wide (e.g., REML 0.692–0.997).

### S5 Excluded Full-Text Studies and Reasons

The following studies were reviewed in full text but excluded from the quantitative synthesis. Reasons for exclusion are summarized below.

- [39] – *AgeML: Age Modeling With Machine Learning*. Reason: Not a diagnostic AD/MCI vs CN study. Notes: Modeled biological/chronological age rather than diagnostic outcomes; no eligible AUC reported. [39]
- [40] – *Digital healthcare for dementia and cognitive impairment: A scoping review*. Reason: Review/scoping review (non-original). Notes: Narrative synthesis only; no patient-level data or extractable metrics.
- [41] – *Artificial intelligence in brain MRI analysis of Alzheimer’s disease over the past 12 years: A systematic review*. Reason: Systematic review (non-original). Notes: Summarizes prior studies; no new data.
- [42] – *Adversarial Network-Based Classification for Alzheimer’s Disease Using Multimodal Brain Images: A Critical Analysis*. Reason: Methods/critique paper. Notes: Discusses adversarial frameworks but lacks standardized diagnostic metrics.

### S6 Certainty of Evidence

Starting at high for diagnostic accuracy, we downgraded for: (1) risk of bias (many internal-only validations; small samples), (2) indirectness (ADNI/OASIS dominance), and (3) publication bias (Egger intercept significant). We upgraded when studies used multi-centre external validation.

Overall certainty: moderate but interpret with caution

